# Precision Electroconvulsive Therapy (PET) project: tackling (cost) effectiveness and patient’s perspectives

**DOI:** 10.1101/2024.03.21.24304665

**Authors:** Philip van Eijndhoven, Indira Tendolkar, Dore Loef, Jordy Rovers, Metten Somers, Harm Pieter Spaans, Jeroen van Waarde, Bart Schut, Ben Wijnen, Esmée Verwijk, Annemieke Dols

## Abstract

**Background:** Depression puts a great burden on both patients, relatives and society as a whole. Electroconvulsive therapy (ECT) is regarded as a safe and effective treatment for severe and chronic depressive episodes, even when other interventions such as psychotherapy or psychopharmacology have failed. Despite its superior efficacy, use of ECT for depressed patients is surprisingly low in most European countries as exemplified in a recent Dutch study. This low application rate is possibly due to (1) limited knowledge on the optimal position of ECT in the treatment algorithms, (2) a lack of knowledge on cost-effectiveness, (3) fear for (cognitive) side-effects in patients, relatives and professionals, and (4) the outdated representation of ECT in the media and society at large.

**Methods:** The present study will overcome the aforementioned limitations and setup a large database of merged clinical and research cohorts of ECT-patients (N=±1500) and an observational prospective cohort study, in order to analyze aspects of (cost-) effectiveness and side-effects of ECT in retro- and prospective Dutch data. Using these results and together with qualitative information from patients and their relatives, we will disseminate the gained knowledge and develop with a decision-making tool that will guide future patients and their family members referred for ECT.

**Discussion:** Our project will further clarify the position of ECT in treatment algorithms for depression based on scientific data, including data on cost-effectiveness, cognitive side-effects and needs of the patients in the decision-making process. By these means, it will contribute to the development of successful personalized treatment and preventive strategies also in other countries in cooperation with stakeholders such as national and international commissions.

## Background

Major depression, when complicated by severe symptoms, recurrence or chronicity, puts a great burden on patients, relatives and society as a whole. Although there are several effective treatment options, approximately one third of patients with depression will not respond to pharmacological and /or psychotherapeutic interventions. A chronic course leads to more individual suffering, more somatic comorbidity and higher suicide rates (Spijker et al., 2002). Electroconvulsive therapy (ECT) offers an effective alternative treatment, that leads to remission rates of 50 to 80%, even so in treatment resistant depression (Dierckx, Heijnen, van den Broek, & Birkenhäger, 2012; Husain, Kevan, Linnell, & Scott, 2004). However, the fear of possible cognitive side-effects is a major barrier for patients, relatives and even some professionals. This overshadows the choice for ECT though the effect sizes of treatments using psychotherapy and medication are smaller, with psychotherapy being relatively costly and pharmacotherapy showing a reduction in treatment response with each new attempt following a failed antidepressant course. Also, adequate knowledge on the exact predictors of response, risk factors for cognitive side-effects and follow-up on these effects is lacking.

In the stepped care model of depression, there is a tendency to regard ECT as a ‘treatment-of-last-resort’, which may have unintentionally contributed to too high barriers to initiate ECT in patients who could potentially benefit. For example, in the Netherlands, it has been shown that only a small percentage (1.2 %) of all patients with chronic depression received ECT (Scheepens et al., 2019). Undertreatment with ECT may be caused by issues at three different levels: to (1) limited knowledge on the optimal position of ECT in the treatment algorithms, (2) a lack of knowledge on cost-effectiveness, (3) fear for (cognitive) side-effects in patients, relatives and professionals, and (4) the outdated representation of ECT in the media and society at large.

In the era of precision medicine and wish for shared-decision making, we need to identify more precisely which patients would most likely benefit from ECT in a relative early phase of disease and which patients are most at risk for cognitive side-effects. Two recent meta-analyses revealed that psychotic features, an older age at presentation, shorter duration of the index-episode and the absence of resistance to medication are indicators of a positive ECT-response (van Diermen et al., 2018, Haq, Sitzmann, Goldman, Maixner, & Mickey, 2015). However, these factors showed collinearity and data on the subject level were missing to control for confounders. It is unclear after how many failed medication trials ECT should be advised, although we do know that chances of remission decrease dramatically after the second medication step (Pigott, Leventhal, Alter, & Boren, 2010). The gold standard to study therapeutic efficacy is a randomized, placebo-controlled trial. Including a sham-ECT condition however, would be unethical, since the superior efficacy of ECT has already been shown. Moreover, randomizing patients to ECT or an additional step of medication was found unfeasible as patients would be unwilling to participate in such a study design (Stek, van der Wurff, Uitdehaag, Beekman, & Hoogendijk, 2007).

Consequently, alternative research designs are needed to analyze if earlier treatment with ECT in the Netherlands will be more effective. Also, it seems important to take mixed methods into account when looking at different aspects of ECT. Therefore, we will thoroughly investigate clinical predictors of ECT-outcome by analyzing archival data collected in prospective as well as retrospective clinical studies. In addition, longitudinal observational clinical data will be used to gain more insight into cognitive side-effects. While archival data may help us identify clinical predictors across different representative treatment sites, it is also important to conduct a prospective clinical trial on potential cost-effectiveness of ECT in comparison to pharmacological treatments. Both approaches are necessary to establish the optimal position of ECT in the treatment algorithms for depression. Finally, it is necessary to better understand the needs of the patients and their relatives in the shared-decision making process for ECT. To the best of our knowledge, our project is the first in the Netherlands and worldwide, to examine the optimal position of an ECT-course within a patient’s antidepressive treatment journey based on real-world data in a high-income country.

## Methods

Our overall aim is to base the optimal position of ECT in the treatment algorithm for depression in guidelines on scientific evidence instead of on consensus, to examine its cost-effectiveness and thereby stimulating resource allocation discussions and its implementation in clinical practice. Moreover we aim to reduce the stigma around ECT, so that ultimately patients and relatives can consider ECT as a valuable treatment option based on facts. To achieve this, we will conduct three different work packages (WP1-3), as explained in detail below.

### WP1: Evaluation of effectiveness and safety of ECT and its predictors

This work package will realize two aims: (1) investigate the effectiveness of ECT and determine its clinical predictors in the Netherlands, (2) investigate cognitive side-effects of ECT and determine its clinical predictors in the Netherlands, (3) determine the optimal position of ECT in the depression treatment algorithm.

### Participants and study design of WP1

A database —the Dutch ECT Cohort (DEC)— will be compiled from available clinical and research cohorts of patients that received ECT for (moderate) severe depressive episodes in the Netherlands during the last 20 years. ECT data will be retrieved from all three sorts of health organizations providing ECT in the Netherlands (i.e., general hospitals [GH], university medical centers [UMC] and institutions for mental health care [MHI]) to gather data representative for the current Dutch ECT practices. Twelve large ECT-centers will provide their coded databases regarding the essential variables for this study. A total of approximately 1500 patients will be included in the database. As has also been approved by the ethical board, patient burden due to this study is absent because archival data will be collected and patients have consented for the original studies. For the patients in the clinical databases, an opt-out was offered in cases where the correct contact information was available upon advice of privacy officers. If active opt-out was not feasible e.g. because of lacking contact information or database size, local privacy officers performed a data protection impact assessment according to GDPR guidelines.

### Data-extraction protocol

About half of the data (n=684) will come from local research cohorts for which complete variables of interest are available (Birkenhager et al, 2019; Dols et al., 2017; Guloksuz et al., 2015; Heijnen et al., 2019; Oudega et al., 2010; Scheepens et al., 2020; van Eijndhoven et al., 2016; van Oostrom et al., 2018; van Waarde et al., 2015; Verwijk et al., 2013). Next, additional data will be derived from clinical cohorts of our national partners as cited above. At these involved sites, patient data have either been stored in prospectively collected clinical databases or in their respective electronic patient files (EPF) which will allow digital data collection for several key parameters (n=1104).

All sites evaluate severity of depression by either systematically applying the Hamilton Depression Rating Scale (HDRS; Hamilton 1967), the Montgomery-Åsberg Depression Rating Scale (MADRS; Montgomery & Åsberg, 1979) or the Inventory for depressive Symptomatology (IDS; Rush et al, 1986). Generally, these scales are administered on a weekly basis during ECT-courses. These scales apply validated cut-off values to determine response, remission or non-response of depression, thus enabling a combined analysis for the respective outcomes. Subitems scores of these scales will be collected as well. Furthermore, described treatment outcome in the discharge letter was also assessed. Table 1 gives an overview of the clinical characteristics that will be included in our database based on previous meta-analyses (Haq et al., 2015; van Diermen et al., 2018, Peeters et al., 2016).

**Table 1.**
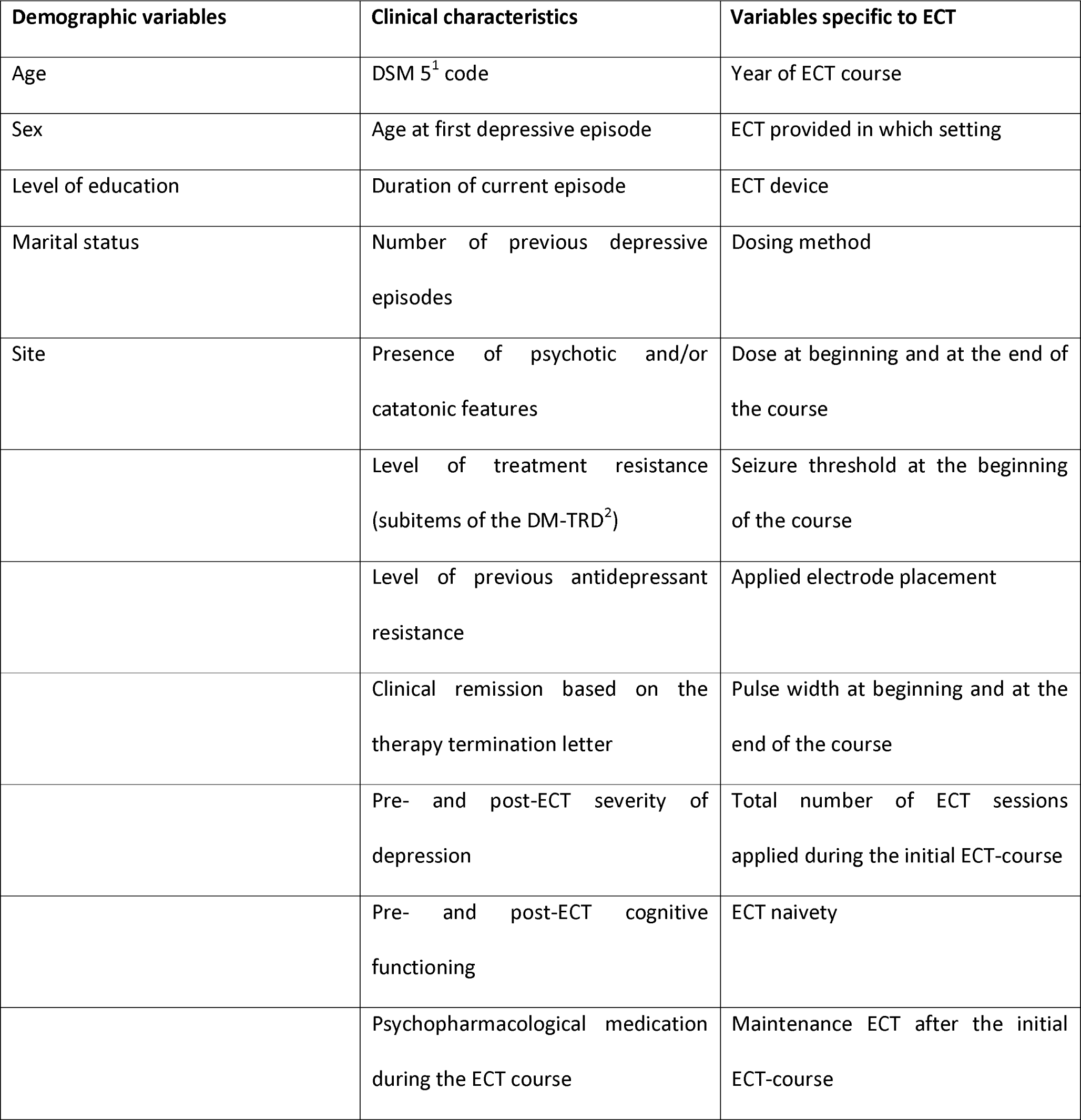
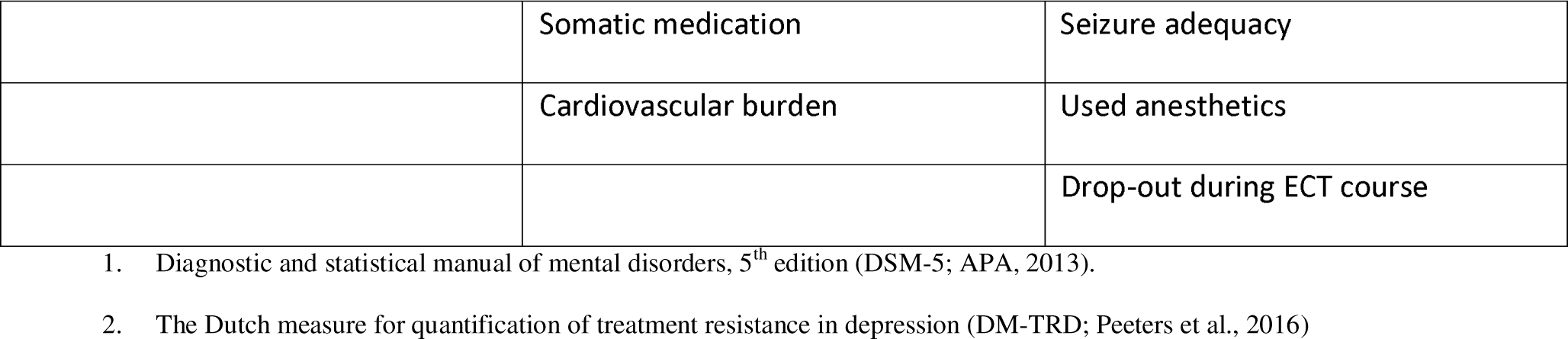
Variables included in the Dutch ECT Cohort.

| Demographic variables | Clinical characteristics | Variables specific to ECT |
| --- | --- | --- |
| Age | DSM 5<sup>1</sup> code | Year of ECT course |
| Sex | Age at first depressive episode | ECT provided in which setting |
| Level of education | Duration of current episode | ECT device |
| Marital status | Number of previous depressive episodes | Dosing method |
| Site | Presence of psychotic and/or catatonic features | Dose at beginning and at the end of the course |
|  | Level of treatment resistance (subitems of the DM-TRD<sup>2</sup>) | Seizure threshold at the beginning of the course |
|  | Level of previous antidepressant resistance | Applied electrode placement |
|  | Clinical remission based on the therapy termination letter | Pulse width at beginning and at the end of the course |
|  | Pre- and post-ECT severity of depression | Total number of ECT sessions applied during the initial ECT-course |
|  | Pre- and post-ECT cognitive functioning | ECT naivety |
|  | Psychopharmacological medication during the ECT course | Maintenance ECT after the initial ECT-course |
|  | Somatic medication | Seizure adequacy |
|  | Cardiovascular burden | Used anesthetics |
|  |  | Drop-out during ECT course |

1. Diagnostic and statistical manual of mental disorders, 5<sup>th</sup> edition (DSM-5; APA, 2013). 2. The Dutch measure for quantification of treatment resistance in depression (DM-TRD; Peeters et al., 2016)

For each subject, only the first course of ECT in the electronic database will be included, preventing enrichment for individuals repeatedly treated with ECT as a result of previous good response to ECT. Several variables specific for ECT will be included for descriptive purposes such as applied electrode placement at the end of the ECT-course (right-unilateral [RUL], bifrontotemporal [BT] of bifrontal [BF]), total number of ECT-sessions applied during the ECT-course, assessment of seizure adequacy during the course (operationalized by the presence of a mean duration of motor (EMG) seizures >= 15 seconds or electroencephalographic (EEG) seizures >= 25 seconds), used anesthetics, and drop-out during the ECT-course due to clinical reasons or side-effects (e.g., headache, muscle aches, postictal delirium, nausea).

### Analyses

Our primary outcome is remission, operationalized as a depression score on the HDRS-17 of 7 or less or on the MADRS of 9 or less at the end of the ECT-course. Our secondary outcomes are response and the presence of ‘clinically significant cognitive side-effects’, operationalized as a broad composite score of the different available test instruments used. Patients’ cognitive performance will be quantified using reliable change index (RCI) methods, to calculate the presence or absence of clinically significant decline in cognitive performance after ECT. A score of RCI = -1.645 is seen as a clinically significant decline (Jacobson & Truax, 1991).

### Aim 1. Investigate the effectiveness of ECT and determine its clinical predictors

We will investigate possible well-known predictors (i.e., psychotic features, an older age, shorter duration of the index episode, absence of medication resistance, age at onset, psychiatric or somatic comorbidities) for remission (primary outcome) and cognitive side-effects (secondary outcome) controlling for confounders (sex) and collinearity. We will also control for referral bias as for example psychotic features are associated with high remission rates and prompt referral for ECT. Primary outcome measures will be on pre-post effects of ECT on depression.

Given the complex nature of the merged data and in line with earlier work from our group members, machine learning approaches will also be used to analyze the data. To obtain the most optimal model, the following approach will be used to analyze the data:

- A priori identification of possible predictors: existing literature and expert opinion will be used to see which predictors likely play an important role in achieving remission and whether additional feature engineering (i.e., creating more informative variables from existing variables) is required.
- Splitting the data in a training- and test-set: The estimated size of the dataset allows us to split into a training-set and a test-(hold-out) set to assess model accuracy. A training-set is a random subset of the data that was used to train the various models. A testing-set is a random subset of the data (mutually exclusive from the training-set) that is used to assess performance of the models on new data.
- Fit various statistical learning models on training-set: Next, different statistical learning techniques will be investigated for their applicability on the data. Common techniques are (linear/logistic) regression, support vector machine models, random forests or neural networks. The different models will be evaluated on their ability to predict remission in patients by cross-validating the models. Cross-validation is a technique to estimate the accuracy of a model by repeatedly partitioning the data and train the data on the training-subset and validate on the other.
- Evaluating the model on the test-set: The model with the best cross-validated performance will be used to predict disability weights for every individual in the testing-set.
- Determine plausibility and clinical relevance with patients and experts: After determining the final model, patient profiles and indicators for early ECT will be drafted in agreement with the various participating experts. These results will also be used in WP3.

### Aim 2. Investigate cognitive side-effects of ECT and determine its clinical predictors

In a subset of the DEC (n=576), more extensive data on cognitive functioning before and after ECT (directly after the course or within the first year) will be available and will be used to 1) examine the evolution of cognitive performance over time following ECT and 2) to identify clinical and demographic predictors for the risk of developing cognitive side-effects following ECT. By calculating norm-scores of the various tests within one of the four main cognitive domains (attention, memory, executive function and language) results of various cohorts on numerous patients will be obtained in more detail with more power. Presence or absence of individual clinically significant cognitive decline or improvement will be operationalized by calculating the reliable change index (RCI, Jacobsen & Truax 1991). Pre- and post-ECT cognitive scores of neuropsychological data will be taken as a starting point for cognitive change.

### Aim 3 Determine the optimal position of ECT in the depression treatment algorithm

For this aim, the effectiveness of ECT ‘earlier’ versus ‘later’ in the patient’s treatment journey will be determined while controlling for possible confounding factors.

The timing of ECT (‘earlier’ vs ‘later’) will be operationalized by categorizing patients by progressive completion of the five steps of the European and Dutch Guideline for Depression (i.e., first selective serotonin reuptake inhibitor [SSRI], second SSRI, tricyclic antidepressant [TCA], lithium addition and irreversible mono-amine oxidase [MAO] inhibitor, respectively) before receiving ECT. Using ECT at an early treatment stage will be defined as having received 0 to 2 adequate pharmacological treatment attempts, while ‘late’ usage will be defined as having undergone > 2 adequate pharmacological treatment courses.

The dichotomization of the DEC cohort in early ECT vs late ECT will allow us to compare the relative efficacy as a function of the level of treatment resistance and inverse propensity weighting will be used to compare these cohorts. Inverse propensity weighting (also known as inverse probability of treatment weighting) is a statistical technique that could substitute for randomization to treatment conditions when the latter is not feasible (Thoemmes & Ong, 2016; Austin, 2008). Based on baseline variables such as age, sex, severity of depression and presence of psychotic symptoms, weights will be estimated to help to make the cohorts comparable with respect to prognostically relevant patient characteristics, and only differing on characteristics related to the treatment resistance and duration of the index period.

Choosing the best set of covariates will be done in a data-driven (empirical) way by checking if baseline balance is achieved after weighing with regard to prognostically relevant variables. The propensity weights will be incorporated in the statistical analysis of outcome parameters by means of inverse propensity score weighted generalized mixed models. This will deliver intention-to-treat (ITT) estimates of efficacy. Logistic regression analysis, including ‘ECT in early treatment stage yes/no’ as predictor and selected variables associated with P-values under 0.10 in the univariate analyses (see Aim 1) will be performed with the primary and secondary outcomes as dependent variables. All tests will be two-sided, with P value set to 0.05 to infer statistical significance. Although these observational data will not permit a conclusion on causal inferences, this design will allow for a comparison of effect-sizes (ES) of early vs late ECT (e.g., early ECT shows a larger ES, both early and late ECT have equal ES, late ECT shows a larger ES).

### WP2: Prospective assessment of (cost)-effectiveness and side-effects of depressive patients treated with either ECT or TCA

For a complete economic evaluation of ECT in the Netherlands, we set two aims: (1) determine cost-effectiveness of providing ECT compared to medication by determining the impact of ECT on remission rates, side-effects and quality of life using A) a one-year time horizon (trial-based economic evaluation) and B) a lifetime time horizon (model-based economic evaluation); and (2) estimate the impact on cost-effectiveness of providing ECT as an earlier versus a later treatment step on remission rates and quality of life.

To determine the cost-effectiveness of ECT compared to medication, a trial-based (prospective) and model-based health economic evaluation will be conducted in accordance with the Dutch guidelines for economic evaluations (Zorginstituut Nederland, 2016). Within both approaches the impact of providing earlier vs later ECT will be integrated. Health economic evaluations will be performed from both the health care system’s perspective as the societal perspective (Zorginstituut Nederland, 2016).

### A. Trial-based economic evaluation in WP2

We will conduct an economic evaluation alongside a multisite, observational prospective cohort study (PROSPECT) with two parallel inception cohorts of patients with major depressive disorder who will be treated with either ECT or TCA. This prospective cohort design is chosen because a randomized controlled trial is not feasible and because it better reflects real-life conditions for comparative cost-effectiveness analyses. Moreover, it is more pragmatic to monitor sustained effectiveness, adverse events and cognitive side-effects during longer follow-up periods after start of ECT or antidepressants, which are important factors to consider for judgements about clinical benefits and cost-effectiveness. The risk of bias is an important limiting factor in observational studies. In this context, ECT may be initiated for more severe conditions (confounding by indication), the cohorts may differ in treatment preferences and adherence to treatment protocol (selection bias), and the cohorts may differ in relapse rates at different times after the initial treatment period (time-varying confounding). To cope with the risk of biases, PROSPECT is setup in accordance with the ROBINS-I tool (http://www.riskofbias.info), specifically designed for studies with a cohort-type of design, in which individuals who are receiving different interventions are followed up over time. The ROBINS-I tool assesses seven domains in which bias might occur in non-randomized intervention studies, and which can be addressed by careful consideration of the risks of bias, systematic assessment of confounding factors, a detailed treatment protocol and explicit reporting guidelines (including a clinical trial registry at clinicaltrials.gov identifier: NCT05306184; Sterne et al., 2016). To test for biases in the observational design, here we will also use inverse propensity weighting to compare the two prospective cohorts (Austin, 2008).

### Data-collection

110 patients (>17 years) with a unipolar MDD who will start ECT and 110 patients (>17 years) with a unipolar MDD who will undergo treatment with antidepressants will be recruited from specialized clinics (i.e., GH, UMC and MHI) and followed-up for one year to gather reliable data on efficacy including relapse rates, cost-effectiveness and quality of life. For an overview of all collected data at the different timepoints see table 2.

**Table 2.** Overview of the variables used in the prospective study (Work package 2)

| Variables | Timeline |  |  |  |  |  |
| --- | --- | --- | --- | --- | --- | --- |
| Clinical variables | T0 | T1 | T2 | T3 | T4 | T5 |
| Hamilton Depression Rating Scale 17 (HDRS-17) | X |  | X |  | X | X |
| Dutch Measure for Therapy Resistance in Depression (DM-TRD) | X |  |  |  |  |  |
| Anti-depressant side-effect checklist (ASEC) | X |  | X |  | X | X |
| Anti-depressant treatment history form (ATHF) | X |  |  |  |  |  |
| Life Chart Interview (LCI) |  |  | X |  | X | X |
| Structured Clinical Interview DSM-5 (SCID-5-S) | X |  |  |  |  |  |
| Cost-effectiveness variables |  |  |  |  |  |  |
| Euro Quality of Life-5D-5L (EQ-5D-5L) | X |  | X |  | X | X |
| The Trimbos Institute and iMTA Cost questionnaire for Psychiatry (TiC-P) | X |  | X |  | X | X |
| Cognitive variables |  |  |  |  |  |  |
| Dutch Adult Reading Test (DART) | X |  |  |  |  |  |
| 15 words test (15WT) | X | X |  | X |  |  |
| Montreal Cognitive Assessment (MOCA) | X | X |  | X |  |  |
| Category and letter fluency | X | X |  | X |  |  |
| Subjective Assessment of Memory Impairment (SAMI) | X | X |  | X | X | X |
| Columbia University Autobiographical Memory Interview (CUAMI) | X | X |  | X |  |  |

T0 = Before start ECT/TCA treatment
T1 = Within a week after the stop with ECT treatment (for the ECT group only)
T2 = After three months
T3 = 12 weeks after the stop with ECT / 20 weeks after the start with TCA
T4 = After 6 months
T5 = After 12 months

The Structured Clinical Interview for DSM 5 disorders (American Psychiatric Association, 2017) will be used to assess diagnosis. The Dutch Method for quantification of Treatment Resistance in Depression (DM-TRD) (Peeters et al., 2016) will assess and quantify the level of treatment resistance. The Hamilton Depression Rating Scale 17 item version (HDRS-17), an observer rated scale to assess the severity of depressive symptoms (Hamilton, 1967), will be assessed to measure the severity of depression.

In keeping with the other work packages, criteria for inclusion in the prospective cohorts will be:

- adult patients (>17 years) with a major depressive disorder who will either start with ECT or TCA;
- failed response to at least one adequate dose-duration trial with antidepressants;
- moderate or severe depression (HDRS-17 >16). Exclusion criteria will be:
- lifetime diagnosis schizophrenia or schizoaffective disorder, current substance abuse disorder, organic brain syndrome;
- the presence of a concurrent significant somatic condition impeding the ability to participate.

In- and exclusion criteria will be checked by psychiatrists from the participating centers. Patients will be enrolled in the groups via usual clinical care. If the patient is eligible, a researcher will provide information for this trial. Within a week, informed consent will be obtained and a personal overview of the study-set-up will be given. We will use CASTOR as a validated data management tool, available for all researchers of the collaborating institutes, and will appoint a monitor from the Clinical Research Centre Nijmegen, Radboudumc, who is certified in Good Clinical Practice and fulfills the requirements for monitoring clinical trials. Inclusion rate will be monitored through https://zorgevaluatienederland.nl/evaluations/prospect

### Intervention(s)

ECT will be applied following a treatment protocol in accordance with the Dutch guideline on ECT for depression (Van den Broek et al., 2021). Treatment with TCA will consist of the protocolled pharmacological treatment step, in accordance with the medication algorithm of the Dutch multidisciplinary guideline for depression (Spijker J, 2013).

### Primary outcome

Depression severity: The Hamilton Depression Rating Scale 17 item version (HDRS-17; Hamilton 1967) after eight weeks of initial treatment and at regular three months intervals in the first year.

### Secondary outcomes

Quality of life: the EuroQol-5D (EQ-5D-5L; Herdman et al., 2011) will be assessed with a validated health-related quality of life (HRQoL) instrument directly after eight weeks of treatment, after three, six, and twelve months. The EQ-5D-5L investigates health states in five different domains: i.e. mobility, self-care, usual activities, pain/discomfort and anxiety/ depression. Current Dutch tariffs will be applied to each of the health states to calculate utilities, anchored at 0 (death) and 1 (full health). Resulting EQ-5D utilities will be merged over time using the trapezium rule resulting in QALY health gains over the entire twelve-month period.

Costs: The Trimbos Institute and iMTA Cost questionnaire for Psychiatry (TiC-P, Bouwmans et al., 2013) will be used to investigate the health care usage and associated costs. The TiC-P is most frequently used for healthcare receipt questionnaire for health-economic evaluation in the Netherlands. Four types of costs will be included: (1) intervention costs (i.e., ECT and TCA), (2) costs stemming from health care utilization, (3) costs for travel and informal care costs of patients and relatives, (4) costs due to loss of productivity (i.e. absenteeism and lesser efficiency in work), in paid work as well as volunteer jobs.

As central clinical endpoint for the cost-effectiveness analysis (CEA), the incremental costs per responder will be used (defined as 50% decrease on the HDRS-17). For the cost-utility analysis (CUA), change in quality of life will be calculated based on the individual pre- and post-treatment quality of life at the initial treatment period and at regular 3 months intervals in the first year.

### Cognitive battery and assessment of side-effects

To measure cognitive functioning, several validated cognitive test instruments will be used to quantify the following cognitive domains: immediate and delayed verbal recall (anterograde memory), autobiographical memory and executive functioning. The tests will be used at baseline, directly after ECT, and at six and twelve months follow-up. Wherever possible, parallel versions are used to prevent learning effects. Tests that will be used are: Dutch Adult Reading Test (DART – premorbid IQ; only at baseline, Schmand et al. 1991); Montreal Cognitive Assessment (MOCA – general cognitive status; all time-points, Nasreddine et al. 2005); Category and letter fluency (language and executive functioning; baseline, 8 weeks and 6 months; Luteijn & Van der Ploeg, 1983; Mulder, Dekker, & Dekker, 2006); 15 woorden-test (verbal learning and memory; baseline, 8 weeks and 6 months; Saan & Deelman, 1986); Subjective Assessment of Memory Impairment (SAMI – subjective memory; baseline, 8 weeks and 6 months; Kumar, Han, Tiller, Loo, & Martin; 2016); Columbia Autobiographical Memory Interview (CUAMI – autobiographical memory; baseline, 8 weeks and 6 months; McElhiney, Moody, Sackeim, Hoogendoorn, & Bergfeld 1997; Verwijk, De Haan, Nwatarali, Hoogendoorn, & Bergfeld, 2021). This test battery takes about 60 minutes at baseline and 45 minutes at eight weeks and six months. The antidepressant side-effects checklist (ASEC; Uher et al., 2009) will be assessed at all timepoints to monitor the side-effects of the TCAs (Hodgson et al., 2015).

### Course of depression

The life chart interview method will be used for detailed information on the course of depression (presence and severity of depressive symptoms) during the year after the start of treatment (after three, six and twelve months) (Lyketsos, Nestadt et al. 1994).

### Sample size calculation

The statistical test will be conducted using a baseline adjusted ANCOVA with four follow-ups (or in an equivalently specified linear mixed model), because this is power-efficient. Taking account of an estimated effect size of 0.3, a correlation between measures of 0.30, we would require N=180 for testing at a=0.05 (2-sided) and a power of (1-ß)=0.80. We will increase sample size to compensate for loss to follow-up (20%) to N=220. The estimated effect size of 0.3 is considered moderate and conservative given that a recent meta-analysis performed by our group, as part of the update of the Dutch guidelines for ECT (van den Broek et al., 2021, demonstrated a standardized mean difference of 0.8 (0.29-1.29) between ECT and antidepressants based on HDRS-scores.

### Analyses

Clinical outcome will be analyzed and reported in accordance with the CONSORT guidelines on an intention-to-treat basis. The sample consisting of all patients will be analyzed using linear mixed modeling. Linear mixed modeling will test intra-group and intergroup differences in effectiveness and side-effects. Level of treatment resistance within the index period will be used as a variable to investigate the effect of early vs late treatment with ECT. Relapse rates during follow up will be analyzed using logistic regression and compared between the two groups. Kaplan-Meier survival estimates will be computed on the basis of the outcome at eight weeks, three, six, nine and twelve months follow-up and the obtained scores will be compared using the log-rank test to account for response rate and time needed to reach 50% reduction of symptoms.

### CEA and CUA: Cost calculations

The total costs will be computed from a bottom-up or micro-costing perspective, with units of health service multiplied by their appropriate unit cost price and added up to deliver an overall total cost estimate (Drummond, Sculpher, Torrance, O’Brien, & Stoddart, 2002). We shall make use of the standard unit cost prices as reported in the latest Dutch guideline for health economic evaluation (Zorginstituut Nederland, 2016, or later editions). Medication costs will be computed with prices based on the Daily Defined Dosage (DDD) taken from www.farmacotherapeutischkompas.nl and www.medicijnkosten.nl, while accounting for the pharmacist’s claw back. Loss of productivity will be assessed with the friction cost method according to the Dutch guideline. Informal care costs will be calculated on the base of shadow prices for unpaid work and the costs of transport will be computed as the mean distance per destination multiplied by standard cost prices. All costs will be expressed in 2018 euros and if needed will be updated to 2018 on the basis of the consumer price index.

The comparability of the groups will be assessed for costs and outcomes, and if necessary, there will be controlled for baseline differences (Manca, Hawkins, & Sculpher, 2005; van Asselt et al., 2009). We will impute missing cost and outcome data with single imputation based on predictive mean matching, and nested in non-parametric bootstraps for intention-to-treat (ITT) analysis (Brand et al., 2019). All cumulative costs and health gains during follow-up will be calculated with “the area under the curve method” to obtain the remission rate and cumulative QALY health gains as accrued over the measurements up to last follow-up at 6 months. The incremental cost-effectiveness ratio (ICER) will deliver the incremental costs per remitter and the incremental costs for each gained QALY. We will perform 5,000 non-parametric bootstraps and plot the simulated ICERs on the ICER plane to account for uncertainty. ICER acceptability curves will be plotted with various willingness-to-pay (WTP) setting to support judgements about whether ECT offers good value for money in comparison to treatment with TCAs. The robustness of findings will be assessed with sensitivity analyses (one-way) directed at uncertainty in the main cost drivers and outcomes (e.g. under different imputation strategies). All research findings will analyzed and reported in accordance with the relevant CONSORT and CHEERS guidelines (Campbell, Piaggio, Elbourne, & Altman, 2012; Husereau et al., 2013; Schulz, Altman, Moher, & the, 2010).

Exploratively, we will examine the impact on cost-effectiveness of providing ECT as an earlier versus a later treatment step on remission rates and quality of life. To explore the impact of early vs. late ECT, an incremental net monetary benefit regression (iNMBR) will be performed. To this extent, a regression model will be constructed to examine associations of treatment-specific characteristics (e.g., early or late ECT) with change net-monetary benefit, while controlling for selected patient-related confounders. Net monetary benefit (NMB) indexes the benefits per patients minus the costs per patient to convert the gain in QALYs into monetary values that express the net benefit.

### B. Model-based health economic evaluation

The model-based health economic evaluation will be conducted according to recommended modelling guidelines (Briggs, Sculpher, & Claxton, 2006; Caro JJ, 2012) and will be informed by both the collected data from the prospective observational multi-site cohort study (PROSPECT) and the DEC cohort, that was described in WP1. The prospective study will inform important parameters not included in DEC such as resource use, relapse and quality of life at different timepoints in the first year. The purpose of the model is twofold: 1) to extend the time horizon of the trial-based health economic evaluation; and 2) to examine usual care in comparison to early or late ECT, given that it is currently considered an ethical challenge to randomize patients to early ECT.

The health-economic simulation will be performed in accordance with modeling techniques proposed by Briggs et al (2006) and follow the recommendations from ISPOR-SMDM Modeling Good Research Practices Task Force-2 (Roberts et al., 2012). The model compares costs between three scenarios: a base-case scenario representing usual / guideline congruent intervention for refractory depression and alternative scenarios representing the early or late intervention with ECT. Cost data will be extracted from the trial.

The long-term costs will be discounted in accordance with Dutch guidelines. The health-economic simulation model will conduct extensive uncertainty analyses overall cost and discounting parameters simultaneously and these will be reported accordingly. Using the model, a budget impact analysis (BIA) will be conducted in accordance with the ISPOR Task Group i.e. Mauskopf et al (2007) and Sullivan et al (2014) to investigate the impact health care budgets in case the new intervention is offered at different implementation levels (Mauskopf et al., 2007; Sullivan et al., 2014). The BIA will be conducted from various perspectives: (1) the perspective of the public purse and (2) the perspective of the health care insurer. For these perspectives the following scenarios will be investigated: a scenario in which ECT is offered to 40%, 60% and 80% of the eligible patients and an extreme scenario in which 100% of these patients will be receiving ECT. This will be done for early- and late-ECT separately to determine the potential increase in healthcare costs when offering more patients ECT, and what the implications are when ECT is offered earlier in the treatment trajectory. The different scenarios will be compared to a base-case scenario where ECT is offered in accordance with the current clinical guidelines for depressive disorder.

### WP3: Qualitative analyses on ECT attitudes and experiences

The first aim of WP3 is to add qualitative analyses to the previously conducted quantitative data analyses about the treatment effects- and side-effects of ECT by conducting qualitative interviews with several stakeholders:

- Patients and their relatives who have received ECT (including positive and negative outcome);
- Patients and their relatives who were referred for ECT and chose not to receive ECT;

**Study design:** Explorative semi-structured interviews in 20 up to 24 patients and their relatives that were treated with ECT or did not choose ECT will be conducted. The ultimate goal is to clarify needs and wishes regarding information on ECT that facilitate shared-decision making. Interviews will also include experienced side-effects in patients that underwent ECT and anticipated side-effects in patients that did not choose for ECT.

## Analyses

The ‘General Interview Guide Approach’ will be used. This approach offers more structure than an informal conversational interview, but also has enough flexibility (Gall, Borg, & Gall, 2003). This means that the interviews will be semi-structured, using a predefined, flexible interview guide with conversation topics and sample open questions. Interview questions will be developed in accordance with the guidelines of McNamara (2009), with open-ended wording, as neutral as possible, asked only once and worded clearly (McNamara, 2009). All interviews are conducted by dedicated team members that have experience in conducting qualitative interviews. The interviews will be audiotaped and transcribed verbatim, coded and stored anonymously. The transcripts of the interviews will be analyzed line-by-line. According to the technique of constant comparative analysis (Grounded Theory), the interview topics will evolve as the interviews progress alongside the data analysis (Charmaz, 2006). In the first stage of data collection, the interviewer will develop a graphical model of the hypothesized links between concepts and categories developed from the data. Through iterative discussion with at least two other researchers, the model will be refined following subsequent interviews. As a quality check, two researchers of the research team will listen to audio-recordings of the first few interviews, in order to analyze in which ways the interviewer’s own views and assumptions may have influenced the posed questions. Validation of the qualitative results will be performed by submitting a questionnaire on the articulated themes to a different sample of ECT-patients, relatives and professionals.

Brief descriptive codes will be assigned to items of data with a focus on aspects that contribute to attitudes towards and experience of ECT and decision-making process to start ECT. The most significant initial codes will be categorized into broader focused codes. These will in turn be analyzed and used to develop categories and subsequently key overarching themes. The final number of participants from the different groups (patients, relatives, professionals) will depend on when ‘meaning saturation’ is reached (Hennink, Kaiser, & Marconi, 2016).

## Discussion

This project will improve knowledge on the indication, application and (cost-) effectiveness of ECT including the burden and likelihood of cognitive side-effects. Stakeholders such as patients and their relatives, clinicians and policymakers should have access to accurate and relevant information upon which healthcare decisions can be based. We will provide this information. With our quantitative (including cost-effectiveness) and qualitative work packages, we will create, enhance and develop knowledge about ECT and its treatment practice in the Netherlands. This will help us in understanding the concerns about ECT and framing them in an evidence-based context on the one hand, while taking biases and stigma into account on the other hand. In collaboration with involved patient organizations, we will raise awareness and expertise for appropriately using ECT in treatment resistant depression that are relevant to other countries as well. We expect the current project to result in a specific knowledge product for cognitive side-effects, because the project will yield a predictive model that may aid the decision and preparation of clinicians and patients by disclosing the expected severity and duration of memory complaints as well as the chance of remission.

Our results will form the basis for future training programs for clinicians, researchers and non-scientific stakeholders to enhance knowledge, optimize clinical applications and improve research on this topic. Lastly, our results will be integrated in a website based decision tool including predictors for ECT success next to additional psychoeducative information for patients that they can use together with their relatives and referring professionals to optimize the process of shared-decision making. From focus groups of the National Dutch Working Group on ECT, we know that it is important to thoroughly weigh the individual’s concerns in respect to ECT but also to increase the health literacy with respect to this topic.

Quality of life is very much impaired in patients with treatment resistant depression. Patients with chronic depression often struggle with feelings of guilt and worthlessness. Knowing more about the course of depression and the treatment with ECT may contribute to dealing with these types of feelings. Social knowledge utilization initiatives both at the patients as well as the professional level may contribute to lowering the stigma that accompanies mental disorders, and thereby improve opportunities for an earlier referral of patients that may need ECT, or broader speaking an earlier selection process on solid grounds. Framing ECT as “the last resort” biases its application, but it is also important to share which kind of patients are less likely to benefit from ECT.

### Strengths and limitations

The project at hand may be seen as a national endeavor of a representative European country and we expect that our conclusions may be widely used. It addresses many open questions regarding ECT and will thus ultimately inform on its clinical use. There is a strong commitment of all partners to conduct this project. The collection of archival data is challenging, but this will ensure a large ‘real-world’ dataset on ECT practice with minimal burden on patients. However, we also have to note some limitations of this project. Firstly, we will not be able to randomize between different groups in the prospective part of this project. Moreover, follow-up of the prospective trial is only one year. On the other hand this was chosen as relevant follow-up duration in close collaboration with the patient union for depression and standard cost-effectiveness measures can be based upon this follow-up period.

## Data Availability

All data produced in the present study are available upon reasonable request to the authors

## Abbreviations

ANCOVA: analysis of covariance
ASEC: antidepressant side-effects checklist
ATHF: anti-depressant treatment history form
BF-ECT: bifrontal ECT
BIA: budget impact analysis
BT-ECT: bifrontotemporal ECT
CEA: cost-effectiveness analysis
CONSORT: Consolidated Standards of Reporting Trials
CHEERS: Consolidated Health Economic Evaluation Reporting Standards
CUA: cost-utility analysis
CUAMI: Columbia Autobiographical Memory Interview
DART: Dutch Adult Reading Test
DDD: Daily Defined Dosage
DEC: Dutch ECT cohort
DM-TRD: Dutch Method for quantification of Treatment Resistance in Depression
ECT: electroconvulsive therapy
EEG: electroencephalogram
EMG: electromyogram
EPF: electronic patient files
ES: effect-size
GH: general hospital
HRQoL: health-related quality of life
EQ-5D-5L: EuroQol-5D
HDRS-17: Hamilton Depression Rating Scale-17 item version
ICER: incremental cost-effectiveness ratio
iMTA: Institute for Medical Technology Assessment
iNMBR: incremental net monetary benefit regression
ITT: intention-to-treat
LCI: Life Chart Interview
MAO: mono-amine oxidase
MHI: mental healthcare institution
MOCA: Montreal Cognitive Assessment
NMB: net monetary benefit
PROSPECT: Prospective assessment of (cost)-effectiveness and side-effects of depressive patients treated with either ECT or TCA
QALY: quality adjusted life year
RCI: reliable change index
RUL-ECT: right-unilateral ECT
SAMI: Subjective Assessment of Memory Impairment
SCID-5: Structured Clinical Interview DSM-5
SSRI: selective serotonin reuptake inhibitor
TCA: tricyclic antidepressant
TiC-P: Trimbos Institute and iMTA Cost questionnaire for Psychiatry
UMC: university medical center
WP: work package
WTP: willingness-to-pay
MADRS: the Montgomery-Åsberg Depression Rating Scale
IDS: Inventory for depressive Symptomatology

## Declarations

## Ethics approval and consent to participate

The present study was evaluated by the Medical Ethics Committee of Amsterdam (under the registration code: 2021.0029, 2021.0557) and the Medical Ethics Committee of RadboudUMC (under the registration code: 2021-13224) which confirmed that the Medical Research Involving Human Subjects Act (WMO) did not apply to the study and that an official approval of this study by our committee was not required. Patients were or will be referred by their health care providers and will provide written informed consent.

## Consent to publish

Not applicable.

## Availability of data and material

Data will be made available upon reasonable request by contacting the first or last author.

## Authors’ Contributions

IT, AD, EV, MS, HPS, JW, BS, BW, JR, DL and PE designed the current proposal, with AD as principal investigator. BW designed the cost-effectiveness analyses. Moreover, AD, PE and EV calculated the required sample size for the second work package. The manuscript draft was written by PE, IT and AD. All authors gave valuable feedback on the manuscript and approved the final version.

## Acknowledgements

The authors thank Dick Veltman and Aartjan Beekman for their helpful comments on the previous version of the grant proposal, and Merijn Eikelenboom and Sanne Hilbers of Amsterdam UMC for their aid in drafting the budget.

## References

American Psychiatric Association. (2013). Diagnostic and statistical manual of mental disorders (DSM-5®). American Psychiatric Pub.

Association, A. P. (2017). SCID-5-S Gestructureerd klinisch interview voor DSM-5 Syndroomstoornissen.

Nederlandse vertaling van Structured Clinical Interview for DSM-5® Disorders– Clinician Version (SCID-5-CV), first edition, en User’s Guide to Structured Clinical Interview for DSM-5® Disorders–Clinician Version (SCID-5-CV), first edition en delen van de Structured Clinical Interview for DSM-5® DisordersResearch Version (SCID-5-RV).. Amsterdam: Boom.

Austin, P.C. (2008). A critical appraisal of propensity-score matching in the medical literature between 1996 and 2003. Stat Med, 27(12), 2037–49.

Birkenhager, T. K., Roos, J., & Kamperman, A. M. (2019). Improvement after two sessions of electroconvulsive therapy predicts final remission in inLpatients with major depression. Acta Psychiatrica Scandinavica, 140(3), 189–195.

Bouwmans, C., De Jong, K., Timman, R., Zijlstra-Vlasveld, M., Van der Feltz-Cornelis, C., Tan, S. S., & Hakkaart-van Roijen, L. (2013). Feasibility, reliability and validity of a questionnaire on healthcare consumption and productivity loss in patients with a psychiatric disorder (TiC-P). BMC Health Services Research, 13(1), 217. doi:10.1186/1472-6963-13-217

Brand, J., van Buuren, S., le Cessie, S., & van den Hout, W. (2019). Combining multiple imputation and bootstrap in the analysis of costLeffectiveness trial data. Statistics in Medicine, 38(2), 210–220

Briggs, A., Sculpher, M., & Claxton, K. (2006). Decision Modelling For Health Economic Evaluation.

Broek, v. d. (2021). Dutch Guideline for ECT (richtlijn ECT). https://www.nvvp.net/stream/richtlijn-elektroconvulsietherapie-2010.pdf

Campbell, M. K., Piaggio, G., Elbourne, D. R., & Altman, D. G. (2012). Consort 2010 statement: extension to cluster randomised trials. Bmj, 345, e5661. doi:10.1136/bmj.e5661

Caro JJ, B. A., Siebert U, et al. (2012). Modeling good research practices - overview: a report of the ISPOR-SMDM Modeling Good Research Practices Task Force-1. Value Health, 5(15), 796–803.

Charmaz, K. (2006). Constructing Grounded Theory: A Practical Guide Through Qualitative Analysis. In (Vol. 1).

Dierckx, B., Heijnen, W. T., van den Broek, W. W., & Birkenhäger, T. K. (2012). Efficacy of electroconvulsive therapy in bipolar versus unipolar major depression: a meta-analysis. Bipolar Disord, 14(2), 146–150. doi:10.1111/j.1399-5618.2012.00997.x

Drummond, M., Sculpher, M., Torrance, G., O’Brien, B., & Stoddart, G. (2002). Methods for The Economic Evaluation of Health Care Programmes (Vol. 54).

van Eijndhoven P, Mulders P, Kwekkeboom L, van Oostrom I, van Beek M, Janzing J, Schene A, Tendolkar I.(2016). Bilateral ECT induces bilateral increases in regional cortical thickness. Transl Psychiatry, 6(8):e874.

Gall, M., Borg, W., & Gall, J. (2003). Educational Research: An Introduction. British Journal of Educational Studies, 32. doi:10.2307/3121583

Guloksuz, S., Arts, B., Walter, S., Drukker, M., Rodriguez, L., Myint, A. M., … & Rutten, B. P. (2015). The impact of electroconvulsive therapy on the tryptophan–kynurenine metabolic pathway. Brain, behavior, and immunity, 48, 48–52.

Hamilton, M. (1967). “Development of a rating scale for primary depressive illness.” Br J Soc Clin Psychol 6(4): 278–296.

Haq, A. U., Sitzmann, A. F., Goldman, M. L., Maixner, D. F., & Mickey, B. J. (2015). Response of depression to electroconvulsive therapy: a meta-analysis of clinical predictors. J Clin Psychiatry, 76(10), 1374–1384. doi:10.4088/JCP.14r09528

Heijnen, W. T., Kamperman, A. M., Tjokrodipo, L. D., Hoogendijk, W. J., van den Broek, W. W., & Birkenhager, T. K. (2019). Influence of age on ECT efficacy in depression and the mediating role of psychomotor retardation and psychotic features. Journal of Psychiatric Research, 109, 41–47.

Hennink, M. M., Kaiser, B. N., & Marconi, V. C. (2016). Code Saturation Versus Meaning Saturation: How Many Interviews Are Enough? Qualitative Health Research, 27(4), 591–608. doi:10.1177/1049732316665344

Herdman, M., Gudex, C., Lloyd, A., Janssen, M. F., Kind, P., Parkin, D., … & Badia, X. (2011). Development and preliminary testing of the new five-level version of EQ-5D (EQ-5D-5L). Quality of life research, 20(10), 1727–1736.

Hodgson, K., Tansey, K. E., Uher, R., Dernovšek, M. Z., Mors, O., Hauser, J., . . . McGuffin, P. (2015). Exploring the role of drug-metabolising enzymes in antidepressant side effects. Psychopharmacology, 232(14), 2609–2617. doi:10.1007/s00213-015-3898-x

Husain, S. S., Kevan, I. M., Linnell, R., & Scott, A. I. (2004). Electroconvulsive therapy in depressive illness that has not responded to drug treatment. J Affect Disord, 83(2-3), 121–126. doi:10.1016/j.jad.2004.05.006

Husereau, D., Drummond, M., Petrou, S., Carswell, C., Moher, D., Greenberg, D., . . . Loder, E. (2013). Consolidated Health Economic Evaluation Reporting Standards (CHEERS) statement. Bmj, 346, f1049. doi:10.1136/bmj.f1049

Jacobson, N. S., & Truax, P. (1991). Clinical significance: A statistical approach to defining meaningful change in psychotherapy research. Journal of Consulting and Clinical Psychology, 59(1), 12–19. doi:10.1037/0022-006X.59.1.12

Kumar, D. R., Han, H. K., Tiller, J., Loo, C. K., & Martin, D. M. (2016). A brief measure for assessing patient perceptions of cognitive side effects after electroconvulsive therapy: the subjective assessment of memory impairment. The Journal of ECT, 32(4), 256–261.

Luteijn, F., & Van der Ploeg, F. (1983). Handleiding groninger intelligentietest (git). Lisse, *the Netherlands*: *Sweitz & Zeitlinger*.

Lyketsos, C. G., et al. (1994). “The Life Chart Interview - a Standardized Method to Describe the Course of Psychopathology.” International Journal of Methods in Psychiatric Research 4(3): 143–155.

Manca, A., Hawkins, N., & Sculpher, M. J. (2005). Estimating mean QALYs in trial-based cost-effectiveness analysis: the importance of controlling for baseline utility. Health Economics, 14(5), 487–496. 10.1002/hec.944

Mauskopf, J. A., Sullivan, S. D., Annemans, L., Caro, J., Mullins, C. D., Nuijten, M., . . . Trueman, P. (2007). Principles of good practice for budget impact analysis: report of the ISPOR Task Force on good research practices--budget impact analysis. Value Health, 10(5), 336–347. doi:10.1111/j.1524-4733.2007.00187.x

McElhiney, M., Moody, B., & Sackeim, H. A. (1997). The Autobiographical Memory Interview - Short Form. New York: New York State Psychiatric Institute.

McNamara, C. (2009). General guidelines for conducting interviews. Retrieved from https://managementhelp.org/businessresearch/interviews.htm

Montgomery SA, Åsberg M. A new depression scale designed to be sensitive to change. The British journal of psychiatry. 1979;134(4):382–9.

Mulder, J., Dekker, P., & Dekker, R. (2006). Handleiding Woord-Fluency Test/Figuur-Fluency Test. Nederland, *Leiden*, *Leiden*: *PITS*.

Nasreddine, Z. S. et al. (2005). The Montreal Cognitive Assessment, MoCA: a brief screening tool for mild cognitive impairment. Journal of the American Geriatrics Society, 53, 695–699.

Oudega, M. L., van Exel, E., Wattjes, M. P., Comijs, H. C., Scheltens, P., Barkhof, F., … & Stek, M. L. (2010). White matter hyperintensities, medial temporal lobe atrophy, cortical atrophy, and response to electroconvulsive therapy in severely depressed elderly patients. The Journal of clinical psychiatry, 71(1), 22435.

Peeters, F. P., Ruhe, H. G., Wichers, M., Abidi, L., Kaub, K., van der Lande, H. J., . . . Schene, A. H. (2016). The Dutch Measure for quantification of Treatment Resistance in Depression (DM-TRD): an extension of the Maudsley Staging Method. J Affect Disord, 205, 365–371. doi:10.1016/j.jad.2016.08.019

Pigott, H. E., Leventhal, A. M., Alter, G. S., & Boren, J. J. (2010). Efficacy and effectiveness of antidepressants: current status of research. Psychother Psychosom, 79(5), 267–279. doi:10.1159/000318293

Roberts, M., Russell, L. B., Paltiel, A. D., Chambers, M., McEwan, P., & Krahn, M. (2012). Conceptualizing a Model: A Report of the ISPOR-SMDM Modeling Good Research Practices Task Force–2. Medical Decision Making, 32(5), 678–689. doi:10.1177/0272989X12454941

Rush AJ, Giles DE, Schlesser MA, Fulton CL, Weissenburger J, Burns C. The Inventory for Depressive Symptomatology (IDS): preliminary findings. Psychiatry Res. 1986 May;18(1):65–87. doi: 10.1016/0165-1781(86)90060-0.

Saan, R. J., & Deelman, B. G. (1986). Dutch version of the Rey auditory verbal learning test. Groningen: Department of Neuropsychology, RUG, provisional manual.

Scheepens, D. S., van Waarde, J. A., Lok, A., Zantvoord, J. B., de Pont, B., Ruhé, H. G., . . . van Wingen, G. A. (2019). [Electroconvulsion therapy for persistent depression in the Netherlands; very low application rate]. Tijdschr Psychiatr, 61(1), 16–21.

Schmand B, Bakker D, Saan R, Louman J. (1991) The Dutch Reading Test for Adults: a measure of premorbid intelligence level]. Tijdschr Gerontol Geriatrie, 22(1), 15–19.

Schulz, K. F., Altman, D. G., Moher, D., & the, C. G. (2010). CONSORT 2010 Statement: updated guidelines for reporting parallel group randomised trials. BMC Medicine, 8(1), 18. doi:10.1186/1741-7015-8-18

Spijker J, B. C., Meeuwissen JAC, Vliet IM van, Emmelkamp PMG, Hermens MLM, Balkom ALJM. (2013). Multidisciplinaire richtlijn Depressie (Derde revisie). Utrecht: Trimbos instituut.

Spijker, J., de Graaf, R., Bijl, R. V., Beekman, A. T., Ormel, J., & Nolen, W. A. (2002). Duration of major depressive episodes in the general population: results from The Netherlands Mental Health Survey and Incidence Study (NEMESIS). Br J Psychiatry, 181, 208–213. doi:10.1192/bjp.181.3.208

Stek, M. L., van der Wurff, F. B., Uitdehaag, B. M., Beekman, A. T., & Hoogendijk, W. J. (2007). ECT in the treatment of depressed elderly: lessons from a terminated clinical trial. Int J Geriatr Psychiatry, 22(10), 1052–1054. doi:10.1002/gps.1800

Sterne, J. A., Hernán, M. A., Reeves, B. C., Savović, J., Berkman, N. D., Viswanathan, M., . . . Higgins, J. P. (2016). ROBINS-I: a tool for assessing risk of bias in non-randomised studies of interventions. Bmj, 355, i4919. doi:10.1136/bmj.i4919

Sullivan, S. D., Mauskopf, J. A., Augustovski, F., Jaime Caro, J., Lee, K. M., Minchin, M., . . . Shau, W. Y. (2014). Budget impact analysis-principles of good practice: report of the ISPOR 2012 Budget Impact Analysis Good Practice II Task Force. Value Health, 17(1), 5–14. doi:10.1016/j.jval.2013.08.2291

Thoemmes, F., & Ong, A. D. (2016). A primer on inverse probability of treatment weighting and marginal structural models. Emerging Adulthood, 4(1), 40–59. doi:10.1177/2167696815621645

Uher, R., Farmer, A., Henigsberg, N., Rietschel, M., Mors, O., Maier, W., … & Aitchison, K. J. (2009). Adverse reactions to antidepressants. The British Journal of Psychiatry, 195(3), 202–210.

van Asselt, A. D., van Mastrigt, G. A., Dirksen, C. D., Arntz, A., Severens, J. L., & Kessels, A. G. (2009). How to deal with cost differences at baseline. Pharmacoeconomics, 27(6), 519–528. doi:10.2165/00019053-200927060-00007

van Diermen, L., van den Ameele, S., Kamperman, A. M., Sabbe, B. C. G., Vermeulen, T., Schrijvers, D., & Birkenhäger, T. K. (2018). Prediction of electroconvulsive therapy response and remission in major depression: meta-analysis. Br J Psychiatry, 212(2), 71–80. doi:10.1192/bjp.2017.28

van Oostrom I, van Eijndhoven P, Butterbrod E, van Beek MH, Janzing J, Donders R, Schene A, Tendolkar I. Decreased Cognitive Functioning After Electroconvulsive Therapy Is Related to Increased Hippocampal Volume: Exploring the Role of Brain Plasticity. J ECT 2018 Jun;34(2):117–123.

van Waarde JA, Scholte HS, van Oudheusden LJ, Verwey B, Denys D, van Wingen GA. A functional MRI marker may predict the outcome of electroconvulsive therapy in severe and treatment-resistant depression. Mol Psychiatry. 2015 May;20(5):609–14.

Verwijk, E., De Haan, R. J., Nwatarali, C., Hoogendoorn, M. L. C., & Bergfeld, I. O. (2021). Columbia University - Autobiographical Memory Interview, Short Form, Nederlandse vertaling, v1.1. Amsterdam: Amsterdam UMC, University of Amsterdam.

Verwijk E, Spaans HP*, Comijs HC, Kok RM, Sienaert P, Bouckaert F, … Kho KH. (2013). Efficacy and cognitive side effects after brief pulse and ultrabrief pulse right unilateral electroconvulsive therapy for major depression: A randomized, double-blind, controlled study. Journal of Clinical Psychiatry, 74(11).

ZorginstituutNederland. (2016). Richtlijn voor het uitvoeren van economische evaluaties in de gezondheidszorg. Retrieved from https://www.zorginstituutnederland.nl/publicaties/publicatie/2016/02/29/richtlijn-voor-het-uitvoeren-van-economische-evaluaties-in-de-gezondheidszorg

